# Dynamic Measures of Glycemia in Relation to Hepatic Features in Adults Without Diabetes

**DOI:** 10.64898/2026.09.25.26364009

**Authors:** Bahar Bakhshi, Naznin Sultana, Matthew Nayor, Honghuang Lin, Jiantao Ma, Devin W. Steenkamp, Michelle T. Long, Nicole L. Spartano, Maura E. Walker

## Abstract

**Background & Aims:** Dysglycemia is associated with increased risk for hepatic features, including steatosis and fibrosis. Yet, the associations of dynamic measures of glycemia with hepatic features among individuals without diabetes remain understudied.

**Methods:** We included participants from the Framingham Heart Study (FHS) Third Generation cohorts without diabetes who underwent assessment for hepatic steatosis (controlled attenuation parameter [CAP]) and fibrosis (liver stiffness measurement ([LSM]), using vibration-controlled transient elastography, had ≥3 days of continuous glucose monitor data (blinded Dexcom G6 Pro CGM) and completed a mixed meal tolerance test, containing a total of 600 kcal, 75 g carbohydrate, 21 g fat, and 29 g protein (MMTT, 2022-2025). We performed multivariable linear and logistic regression to investigate the associations of standardized 2-h postprandial glucose (2-h PPG) following MMTT, as well as CGM-derived measures, with CAP and log-transformed LSM, as well as hepatic steatosis (CAP≥274ndB/m) and fibrosis (LSM≥8.2 kPa), adjusting for confounders.

**Results:** In 1071 FHS participants (58.3% female, 56.4% normoglycemia), mean age was 60.2 years and BMI 28.2 kg/m^2^. Higher 2-h MMTT-PPG and CGM summary measures of glycemic burden (e.g., % time >140 mg/dL) were associated with 23-36% higher odds of steatosis and higher CAP. Similarly, CGM summary measures of glycemic burden (e.g., % time >140 mg/dL) and glycemic variability (e.g., coefficient of variation) were associated with 28-59% higher odds of hepatic fibrosis but not log-LSM.

**Conclusion:** Measures of postprandial and continuous glucose dynamics are associated with hepatic features among individuals without diabetes.

## Introduction

Metabolic dysfunction-associated steatotic liver disease (MASLD) is a multisystem condition encompassing a spectrum of pathologic and imaging features, from isolated steatosis to steatohepatitis, advanced fibrosis, and ultimately, cirrhosis.(1) The clinical consequences of MASLD extend well beyond liver-related complications, with extensive evidence highlighting a bidirectional link between MASLD and progression of dysglycemia, increased cardiovascular disease (CVD) incidence, as well as CVD related mortality.(2) While such associations reflect shared pathophysiologic processes, including insulin resistance, MASLD exhibits heterogeneity concerning both hepatic pathology and extrahepatic cardiometabolic outcomes.(3) Recent studies have leveraged established clinical measures to reveal distinct phenotypes characterized by differing disease trajectories, with the goal of enhancing MASLD’s prevention and screening strategies.(3) However, capturing early metabolic dysregulation contributing to hepatic features, particularly in those without prevalent cardiometabolic disease, remains challenging and complementary approaches for early identification prior to progression of hepatic features are warranted.(2)

Wearable devices may offer an opportunity to address this gap via continuous physiologic phenotyping of individuals under free-living conditions.(4, 5) For instance, recent advances in continuous glucose monitoring (CGM) enable high-resolution characterization of glycemic physiology that may otherwise remain undetected by conventional measures (FPG/HbA1c).(6–8) Several aspects of glucose homeostasis, including postprandial glucose exposure and glycemic variability, could hold information relevant to hepatic pathology.(7) Therefore, methods uncovering the postprandial state and continuous glucose dynamics may offer complementary metabolic phenotyping and provide an avenue for early identification of at-risk individuals and risk stratification of those with hepatic features.(9) Studies in individuals without type 2 diabetes (T2D) have shown significant correlations of CGM-derived summary measures with liver attenuation, ultrasound metrics reflecting hepatic steatosis.(10, 11) However, relations of postprandial glucose (PPG) and CGM summary measures with hepatic steatosis and fibrosis among individuals without T2D remains understudied.

Here, among participants of the Framingham Heart Study (FHS) free of T2D, we leveraged 2-h PPG following a mixed-meal challenge, in addition to summary measures derived from CGM, and investigated their associations with hepatic features, including steatosis and fibrosis, measured via vibration-controlled transient elastography (VCTE).

## METHODS

### Study Sample

Participants attending the fourth examination cycle of the FHS Third Generation, New Offspring Spouse, and Omni 2 Cohorts (12) (n=2718) were invited to undergo an oral mixed meal tolerance test (MMTT) and subsequently wear a blinded Dexcom G6 Pro CGM. Of those, we included individuals who had ≥3 days of CGM data, had corresponding 2-h PPG following MMTT, and underwent VCTE from their third examination cycle FHS exam. We excluded individuals according to the steps detailed in **Figure S1**, including those with T2D defined as HbA1c ≥6.5% or FPG ≥126 mg/dL or taking glucose-lowering medications (n=223) and those reporting high alcohol intakes at the time of their VCTE examination (>14 drinks/week for women and >21 drinks/week for men, n=109), resulting in a final sample size of 1071.

All FHS study protocols and procedures were conducted according to the guidelines of the Declaration of Helsinki and approved by the institutional review board for human research at Boston University Medical Campus. All participants provided written consent prior to participation in the study.

### Glycemic Traits

#### CGM summary measures

Participants were asked to wear a blinded Dexcom G6 Pro monitor on their arm or abdomen for up to 10 days. Interstitial glucose values were measured every 5 minutes, providing a maximum of 288 readings/day for the entire wear time, with an average wear time of 8.2 days in the current study sample. CGM-derived summary measures were calculated using the “iglu” R package, a standardized, open-source tool for analyzing and interpreting CGM data.(13) We focused on 10 CGM summary measures capturing complementary aspects of glycemic burden and variability, including mean glucose, % time >140 mg/dL, coefficient of variation (CV), mean amplitude of glycemic excursions (MAGE), mean of daily differences (MODD), continuous overall net glycemic action over 1-hour (CONGA-1), glycemic risk assessment diabetes equation (GRADE), J-index, low blood glucose index (LBGI) and high blood glucose index (HBGI). (10, 14) Each summary measure provides distinct information on glucose dynamics, with detailed description provided in **Table S1**.

#### HbA1c and 2h-PPG following on MMTT

After a 10-hour overnight fast and prior to CGM placement, HbA1c was measured in whole blood samples using standard clinical assays and expressed as a percentage. Participants then consumed a standardized liquid meal containing a total of 600 kcal, 75g carbohydrates, 21g fat, 29g protein within 5 minutes. To measure FPG and 2-h MMTT-PPG, we collected plasma samples prior and 2-h following the challenge.

### Liver Enzyme and Imaging Measurements

VCTE (FibroScan; Echosens, Paris, France) was performed to assess hepatic steatosis (controlled attenuation parameter [CAP]) and fibrosis (liver fibrosis measurement [LSM]) during the third examination cycle of the FHS Third Generation and Omni 2 Cohorts (2016–2019).(15, 16) Prior to the examination, participants fasted for ≥3 hours, and were placed in the supine position with the right arm in maximal abduction, with the VCTE probe positioned in the intercostal space over the right lobe of the liver. For each participant, a minimum of 10 measurements were collected, median values were recorded and were reviewed by a hepatologist (M.T.L.) to ensure adequate imaging quality. In this analytical sample, no participant had poor quality scans with high measurement variability (defined as an interquartile range/median ratio >0.30 with LSM ≥7.1 kPa).(17) To define hepatic steatosis and fibrosis, we used previously determined thresholds of CAP ≥274 dB/m and LSM ≥8.2 kPa, respectively.(17)

At both exam 3 and 4, liver enzymes, including aspartate aminotransferase (AST) and alanine aminotransferase (ALT) were measured following a 10-hour fast, using standardized clinical assays at both examination cycles.

### Genetic Risk Score (GRS)

Among a subset of individuals with available genetic data (n=968), we derived a weighted GRS established in previous studies according to 17 single nucleotide polymorphisms (SNPs) associated with hepatic steatosis.(18, 19) Genotyping was performed with the Affymetrix 550k Array and imputed to the 1000 Genomes Project reference panel. For GRS calculation, SNPs with imputation quality R^2^ > 0.5 and minor allele frequency >0.005 were selected, with the cumulative GRS computed as the sum of each weighted allele using the previously published effect size (GOLD-weighted approach).(18, 19)

### Statistical Analysis

The characteristics of participants according to glycemic status (**Table 1**), as well as prevalent hepatic steatosis (**Table S2**), are reported as mean (SD) for continuous variables and n (%) for categorical variables. We characterized glycemic status as either having normoglycemia (FPG <100 mg/dL and HbA1c <5.7%) or prediabetes (FPG 100-125 mg/dL or HbA1c 5.7-6.4%).

**Table 1.**
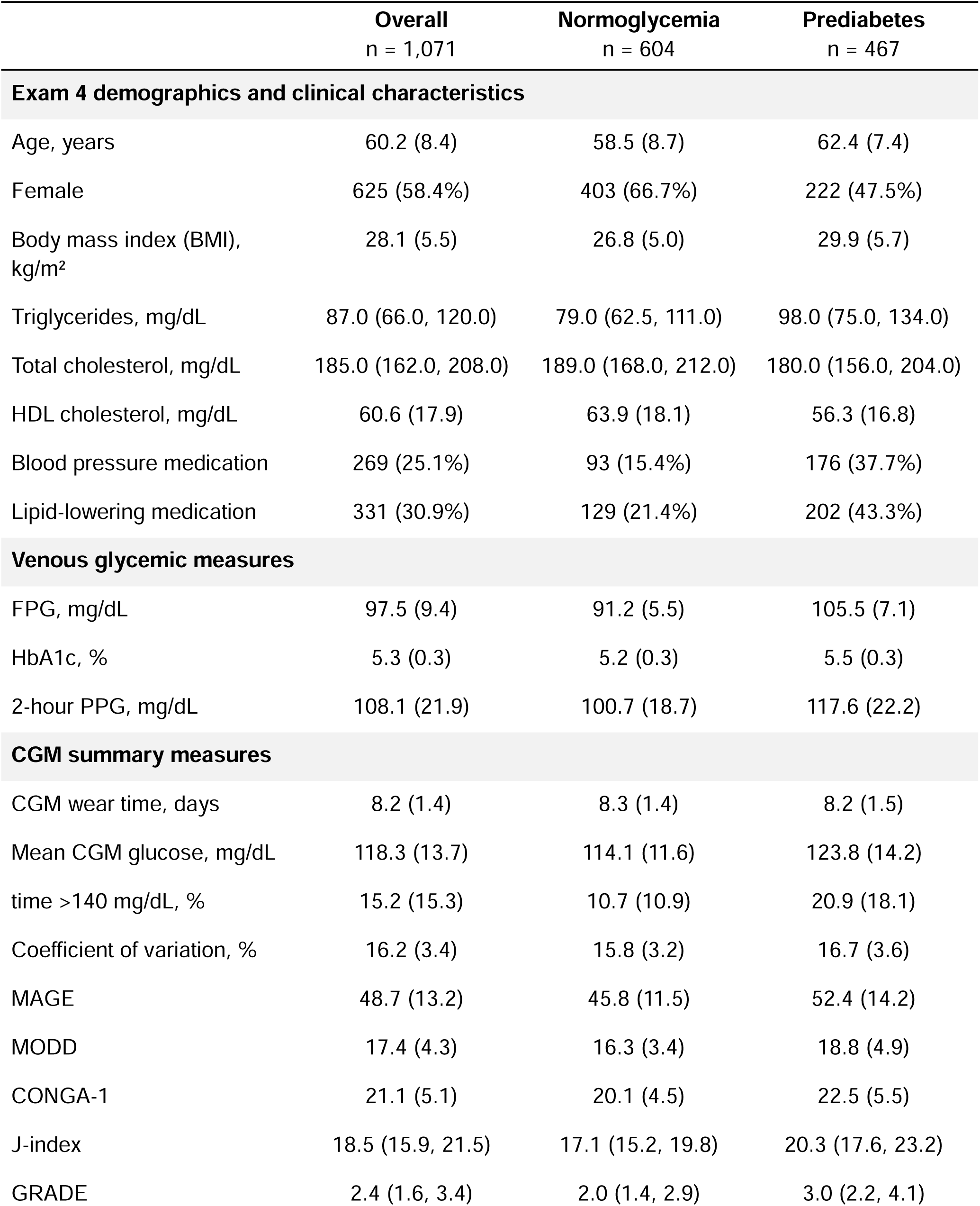
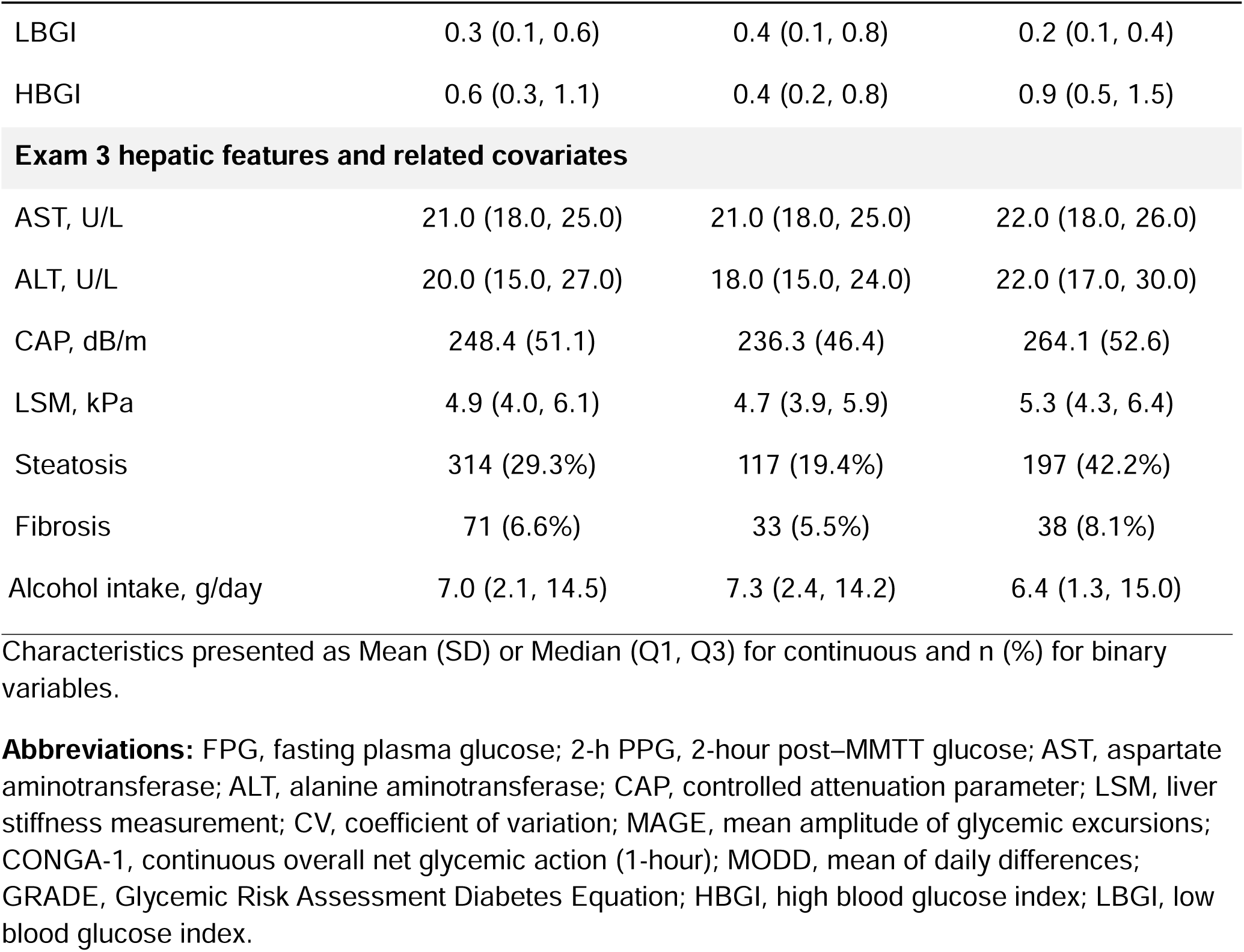
Participant characteristics according to Exam 4 glycemic status.

|  | **Overall**<br>n = 1,071 | **Normoglycemia**<br>n = 604 | **Prediabetes**<br>n = 467 |
| --- | --- | --- | --- |
| **Exam 4 demographics and clinical characteristics** |  |  |  |
| Age, years | 60.2 (8.4) | 58.5 (8.7) | 62.4 (7.4) |
| Female | 625 (58.4%) | 403 (66.7%) | 222 (47.5%) |
| Body mass index (BMI),<br>kg/m<sup>2</sup> | 28.1 (5.5) | 26.8 (5.0) | 29.9 (5.7) |
| Triglycerides, mg/dL | 87.0 (66.0, 120.0) | 79.0 (62.5, 111.0) | 98.0 (75.0, 134.0) |
| Total cholesterol, mg/dL | 185.0 (162.0, 208.0) | 189.0 (168.0, 212.0) | 180.0 (156.0, 204.0) |
| HDL cholesterol, mg/dL | 60.6 (17.9) | 63.9 (18.1) | 56.3 (16.8) |
| Blood pressure medication | 269 (25.1%) | 93 (15.4%) | 176 (37.7%) |
| Lipid-lowering medication | 331 (30.9%) | 129 (21.4%) | 202 (43.3%) |
| **Venous glycemic measures** |  |  |  |
| FPG, mg/dL | 97.5 (9.4) | 91.2 (5.5) | 105.5 (7.1) |
| HbA1c, % | 5.3 (0.3) | 5.2 (0.3) | 5.5 (0.3) |
| 2-hour PPG, mg/dL | 108.1 (21.9) | 100.7 (18.7) | 117.6 (22.2) |
| **CGM summary measures** |  |  |  |
| CGM wear time, days | 8.2 (1.4) | 8.3 (1.4) | 8.2 (1.5) |
| Mean CGM glucose, mg/dL | 118.3 (13.7) | 114.1 (11.6) | 123.8 (14.2) |
| time >140 mg/dL, % | 15.2 (15.3) | 10.7 (10.9) | 20.9 (18.1) |
| Coefficient of variation, % | 16.2 (3.4) | 15.8 (3.2) | 16.7 (3.6) |
| MAGE | 48.7 (13.2) | 45.8 (11.5) | 52.4 (14.2) |
| MODD | 17.4 (4.3) | 16.3 (3.4) | 18.8 (4.9) |
| CONGA-1 | 21.1 (5.1) | 20.1 (4.5) | 22.5 (5.5) |
| J-index | 18.5 (15.9, 21.5) | 17.1 (15.2, 19.8) | 20.3 (17.6, 23.2) |
| GRADE | 2.4 (1.6, 3.4) | 2.0 (1.4, 2.9) | 3.0 (2.2, 4.1) |

|  | **Overall**<br>n = 1,071 | **Normoglycemia**<br>n = 604 | **Prediabetes**<br>n = 467 |
| --- | --- | --- | --- |
| LBGI | 0.3 (0.1, 0.6) | 0.4 (0.1, 0.8) | 0.2 (0.1, 0.4) |
| HBGI | 0.6 (0.3, 1.1) | 0.4 (0.2, 0.8) | 0.9 (0.5, 1.5) |
| **Exam 3 hepatic features and related covariates** |  |  |  |
| AST, U/L | 21.0 (18.0, 25.0) | 21.0 (18.0, 25.0) | 22.0 (18.0, 26.0) |
| ALT, U/L | 20.0 (15.0, 27.0) | 18.0 (15.0, 24.0) | 22.0 (17.0, 30.0) |
| CAP, dB/m | 248.4 (51.1) | 236.3 (46.4) | 264.1 (52.6) |
| LSM, kPa | 4.9 (4.0, 6.1) | 4.7 (3.9, 5.9) | 5.3 (4.3, 6.4) |
| Steatosis | 314 (29.3%) | 117 (19.4%) | 197 (42.2%) |
| Fibrosis | 71 (6.6%) | 33 (5.5%) | 38 (8.1%) |
| Alcohol intake, g/day | 7.0 (2.1, 14.5) | 7.3 (2.4, 14.2) | 6.4 (1.3, 15.0) |

Characteristics presented as Mean (SD) or Median (Q1, Q3) for continuous and n (%) for binary variables.
**Abbreviations:** FPG, fasting plasma glucose; 2-h PPG, 2-hour post–MMTT glucose; AST, aspartate aminotransferase; ALT, alanine aminotransferase; CAP, controlled attenuation parameter; LSM, liver stiffness measurement; CV, coefficient of variation; MAGE, mean amplitude of glycemic excursions; CONGA-1, continuous overall net glycemic action (1-hour); MODD, mean of daily differences; GRADE, Glycemic Risk Assessment Diabetes Equation; HBGI, high blood glucose index; LBGI, low blood glucose index.

#### Primary Analyses

To handle potential outliers and facilitate comparison across predictors with varying scales, CGM measures were winsorized (1^st^ and 99^th^ percentile) and all glycemic traits (HbA1c, 2 MMTT-PPG, CGM summary measures) were standardized (mean = 0 and SD = 1). We performed multivariable logistic regression to investigate the associations of each glycemic trait with hepatic steatosis (CAP ≥274 dB/m) and fibrosis (LSM ≥8.2 kPa) as binary outcomes, in addition to multivariable linear regression to investigate the associations of each glycemic trait with CAP and log-transformed LSM, as continuous outcomes. To address the time lag between data collection periods, we adjusted models for relevant demographic and glycemic covariates at Exam 4, aligning with CGM data collection timeline, in addition to liver-related covariates at Exam 3, parallel with VCTE data collection timeline. Model adjustments included Exam 4 age, sex, smoking, lipid-lowering medication, Exam 3 alcohol intake (Model 1), with additional adjustments for Exam 4 FPG (Model 2), Exam 4 BMI (Model 3), and lastly, Exam 3 AST:ALT ratio (Model 4).

#### Secondary and sensitivity analyses

To investigate the potential modifying effect of GRS on the association of glycemic traits with hepatic features, we tested the interaction term with the continuous GRS and stratified the associations according to top tertile of GRS.

To address the potential confounding impact of transitions in glycemic status between Exam 3 to Exam 4, we first repeated the analyses excluding individuals who had normoglycemia at Exam 3 and progressed to prediabetes at Exam 4 (n=213), and vice versa (n=59). Among those with stable glycemic status across Exam 3 and Exam 4 (i.e., both having normoglycemia at Exam 3 and Exam 4 or both having prediabetes at Exam 3 and Exam 4 [n=803]), we then conducted analyses testing an interaction term for glycemic status and stratifying the associations according to stable normoglycemia and prediabetes.

To address the potential bias of CGM wear duration, we additionally performed a sensitivity analysis excluding participants with <7 days of CGM data (n=186). This resulted in a sample of 885 individuals with ≥7 days of CGM data, in whom we repeated the regression analysis of glycemic traits and hepatic steatosis and fibrosis. To account for multiple testing, we adjusted all the p-values and p-interactions using the Benjamini–Hochberg false discovery rate (FDR) procedure. An FDR-adjusted q-value <0.05 was considered statistically significant. All analyses were performed R/RStudio version 4.5.2.

## RESULTS

In this analytical sample of 1071 FHS participants, 57.4% were female, 55.9% had normoglycemia, with an average age of 59.8 years and BMI 28.1 kg/m^2^ (**Table 1**). Both conventional glycemic metrics (FPG/HbA1c) and CGM summary measures were higher in those with prediabetes compared with normoglycemia. Steatosis prevalence was 29.3% in the overall population, 19.4% in those with normoglycemia, and 42.2% in those with prediabetes. Fibrosis prevalence was 6.5% in the overall population, 5.5% in those with normoglycemia, and 8.1% in those with prediabetes.

### Glycemic Traits and Hepatic Steatosis

Using multivariable logistic regression models, we observed significant associations of glycemic traits with hepatic steatosis in the overall study sample (**Figure 1A, Table S3**) Higher 2-h MMTT-PPG, CGM measures of glycemic burden and composite scores of glycemic control were associated with higher odds of hepatic steatosis, independent of relevant demographic and clinical covariates. For instance, a 1 SD higher 2-h MMTT-PPG (21.9 mg/dL) and % time >140 mg/dL (15.3%) were associated with 36% and 31% higher odds of steatosis, even after adjusting for FPG, BMI, and AST to ALT ratio. However, most CGM measures of glycemic variability were not associated with odds of steatosis. While 1 SD higher MODD was associated with 34% higher odds of steatosis, after subsequent adjustments for FPG, the association was no longer significant. Similarly, while higher HbA1c was associated with higher odds of steatosis, but the association attenuated towards null after further adjustments for BMI.

**Figure 1.**
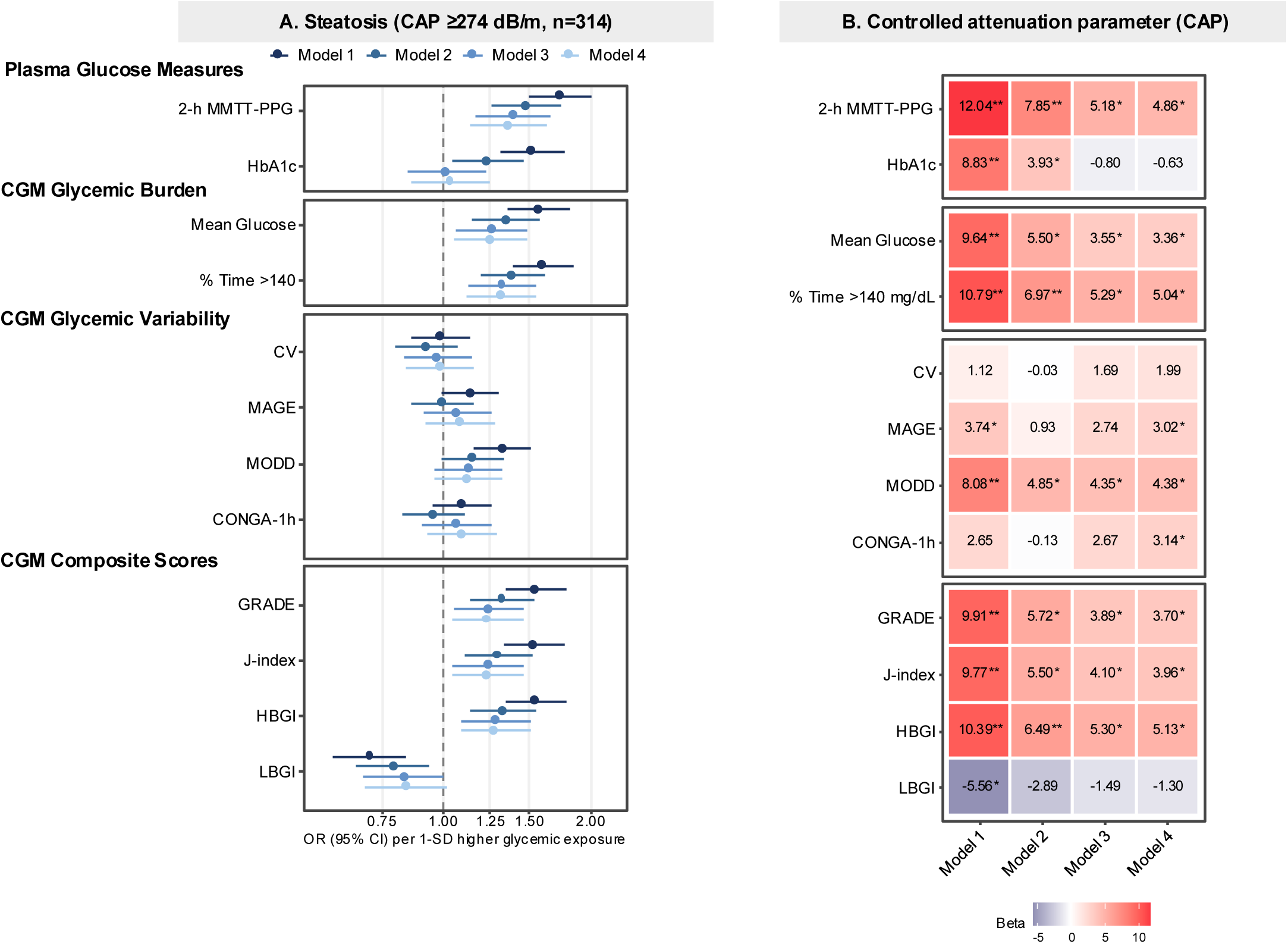
**A.** Multivariable logistic regression models depicting the associations between standardized glycemic traits and hepatic steatosis. **B.** Multivariable linear regression models depicting the associations between standardized glycemic traits and controlled attenuation parameter (CAP). *, ** indicate FDR-corrected p <0.05, <0.001. Model 1 was adjusted for age, sex, smoking, lipid-lowering medication, alcohol intake, model 2 included model 1 with additional adjustment for FPG, model 3 included model 2 with additional adjustment for BMI, model 4: model 3 with additional adjustment for additional adjustment for AST:ALT ratio at Exam 3.

In multivariable linear regression models, we observed significant associations of various glycemic traits with continuous CAP in the overall study sample (**Figure 1B, Table S2**). Higher 2-h MMTT-PPG, CGM measures of glycemic burden and composite scores of glycemic control were associated with 9.64-12.04 dB/m higher CAP, after adjusting for age, sex, smoking, lipid-lowering medication, and Exam 3 alcohol intake. After further adjustment for FPG and BMI, the positive associations of glycemic traits (except for LBGI) with CAP persisted. Additional adjustment for Exam 3 AST to ALT ratio did not substantially attenuate the associations. While most CGM measures of glycemic variability were not associated with CAP, higher MODD was associated with 4.90 dB/m higher CAP, with associations persisting after additional adjustments for FPG, BMI, and Exam 3 AST to ALT ratio

### Glycemic Traits and Hepatic Fibrosis

In multivariable logistic regression models, while 2-h MMTT-PPG and HbA1c were not associated with odds of hepatic fibrosis, higher CGM measures of glycemic burden (i.e., mean glucose), glycemic composite scores of glycemic control (except for LBGI) were associated with higher odds of hepatic fibrosis (**Figure 2A, Table S5**). For instance, 1 SD higher mean glucose mg/dL was associated with 37% higher odds of fibrosis even after FPG, BMI, and AST to ALT ratio. Notably, higher CGM measures of glycemic variability were also associated with higher odds of hepatic fibrosis, with the associations persisting across fully adjusted models. For example, 1 SD higher MAGE (13.2 mg/dL) and MODD (4.3 mg/dL) were associated with 54% and 46% higher odds of fibrosis. However, multivariable linear regression models, glycemic traits were not associated with log-transformed LSM across all models (**Figure 2B, Table S2**). We observed a positive association of higher 2-h MMTT-PPG and log-LSM although with further adjustment for FPG, the association attenuated towards null.

**Figure 2.**
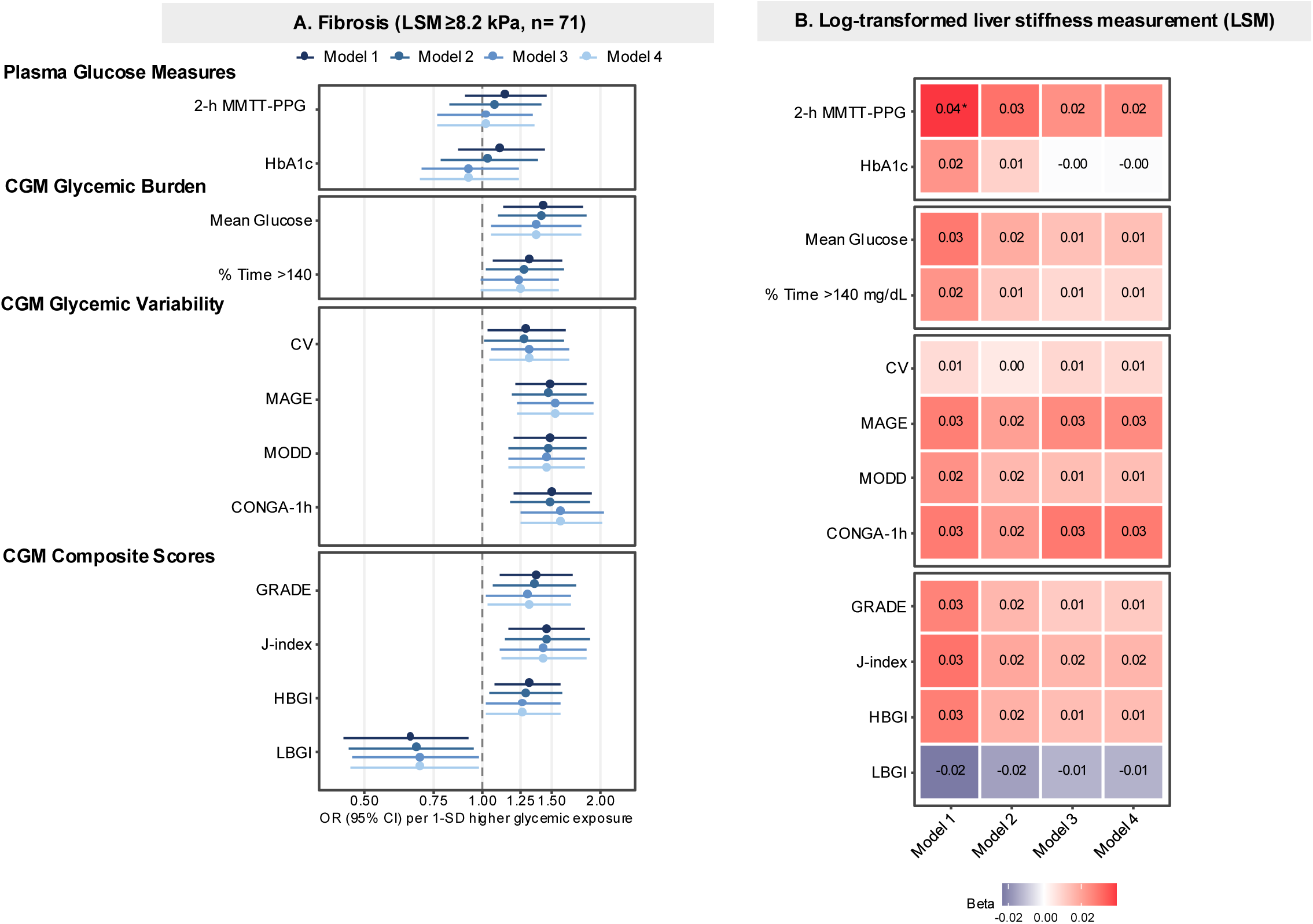
**A.** Multivariable logistic regression models depicting the associations between standardized glycemic traits and hepatic fibrosis. **B.** Multivariable linear regression models depicting the associations between standardized glycemic traits and log-transformed liver stiffness measurement (LSM). *, ** indicate FDR-corrected p <0.05, <0.001. Model 1 was adjusted for age, sex, smoking, lipid-lowering medication, alcohol intake, model 2 included model 1 with additional adjustment for FPG, model 3 included model 2 with additional adjustment for BMI, model 4: model 3 with additional adjustment for additional adjustment for AST:ALT ratio at Exam 3.

### Secondary and Sensitivity Analysis

The interactions between glycemic traits and GRS in relation to hepatic features were not statistically significant (all FDR-corrected p-interactions >0.05). Nonetheless, when stratified by the top tertile of GRS, we did not observe a statistically significant effect modification on the associations between glycemic traits and hepatic features among those in the top tertile or lower two tertiles PRS (**Figure S2** and **S3**).

Among the individuals with stable glycemic status across Exam 3 and Exam 4 (i.e., having normoglycemia at Exam 3 and Exam 4 [n=545] or having prediabetes at Exam 3 and Exam 4 [n=254]), we observed similar associations between glycemic traits and hepatic features in terms of direction and magnitude (**Figure S4 and S5**). The interactions between glycemic traits and glycemic status in relation to hepatic features were not statistically significant (all FDR-corrected p-interactions >0.05). when stratified by glycemic status, we found overall similar associations between glycemic traits and hepatic features among those with normoglycemia and prediabetes (**Figure S6** and **S7**).

When excluding <7 days of CGM wear (**Figure S8**), we observed similar associations between glycemic traits and hepatic features in terms of direction of magnitude to those reported in Figure 1-2.

## DISCUSSION

Among individuals without T2D, we found that glycemic traits reflective of postprandial glucose handling and continuous glucose dynamics are associated with previously measured hepatic features, after considering FPG, BMI, and AST:ALT ratio. While higher 2-h MMTT-PPG, CGM measures of glycemic burden (e.g., mean glucose and % time >140 mg/dL) and composite scores of glycemic control (e.g., J-index and GRADE) were associated with 24-32% higher odds of hepatic steatosis, CGM measures of glycemic variability (e.g., CV and MAGE) were less consistently associated with hepatic steatosis. Additionally, higher CGM measures of glycemic burden and variability were associated with 28–59% higher odds of hepatic fibrosis, but not with log-LSM.

Despite a limited body of evidence, our findings mostly corroborate with studies relating CGM summary measures and hepatic features.(9, 11, 20) An earlier large-scale cross-sectional study of individuals without T2D reported significant age- and sex-adjusted correlations of CGM summary measures (e.g., J-index, MODD, % time >140 mg/dL) with hepatic attenuation, a quantitative ultrasound metric reflecting hepatic steatosis(9, 20) However, age- and sex-adjusted correlations of CGM summary measures and hepatic features remain prone to residual confounding by other key lifestyle and clinical factors. A more recent analysis of the same cohort revealed that over approximately 2.6 years of follow-up, higher % time >180 mg/dL and % time >140 mg/dL were associated with an elevated risk for developing metabolic disease, a composite outcome including dyslipidemia, prediabetes, T2D, and MASLD, although MASLD-specific risk estimates were not statistically significant likely due to a low incidence.(9) Another cross-sectional study of Chinese adults with and without T2D showed that CGM measures of glycemic burden and variability were positively associated with hepatic steatosis severity determined via magnetic resonance imaging, independent of lifestyle and clinical confounders such as BMI.^9^ However, when stratified by HbA1c categories, the associations were only significant among individuals with prediabetes (HbA1c 5.7-6.4%) but not normoglycemia (HbA1c <5.7%).^9^

Our results suggesting a positive association between 2-h MMTT-PPG and hepatic steatosis are further aligned with substantial evidence linking 1-h or 2-h post-load glucose following an oral glucose tolerance test (OGTT) to hepatic features.(21–24) A recent cross-sectional study of individuals with obesity showed that elevated 1-h OGTT-PPG was associated with 3-times the odds of having biopsy-proven MASLD, in addition to significantly higher levels of histological features, such as hepatic steatosis, fibrosis, and lobular inflammation scores.(21) Additionally, an earlier prospective cohort study of Chinese adults without T2D indicated that over approximately 4.4 years of follow-up, higher 2-h OGTT-PPG was associated with higher incidence of ultrasound-defined hepatic steatosis, as well as advanced fibrosis.(24) While in our study, the mixed meal challenge contained three main macronutrients to resemble a typical real-life meal, the total amount of carbohydrates was matched to that of an OGTT (75 g), making it well suited for investigating postprandial glucose handling in a more physiologically relevant manner.(25)

We additionally observed that HbA1c was not associated with the odds of hepatic steatosis or fibrosis after adjusting for BMI, and AST:ALT ratio, among individuals without T2D. Although substantial longitudinal evidence has linked HbA1c as a marker of chronic glycemia, to hepatic features particularly in those with T2D, our findings underscore the potential importance of utilizing novel approaches to characterize postprandial glycemia and temporal glycemic states for identifying subclinical perturbations in glucose homeostasis, and therefore, early or intermittent metabolic dysregulation that may concomitantly inform hepatic features.

To address the heterogeneity across MASLD development and progression, recent studies have employed data-driven clustering approaches on established clinical measures and identified two major phenotypic profiles, a cardiometabolic and liver-specific, with distinct prognosis as well as long-term hepatic- and extrahepatic features.(26, 27) Importantly, the cardiometabolic cluster exhibited pronounced dysglycemia and was subsequently associated with higher risk for developing T2D and CVD, highlighting the importance of timely diagnosis and early interventions to manage the cardiometabolic conditions underlying hepatic features.(3). (26, 27) Future studies are needed to investigate the potential added value of utilizing CGMs in identifying clinically relevant subtypes of hepatic and, as well as risk stratification of hepatic features, among those with and without T2D.

Our study has several strengths and limitations. We included a robust sample of individuals with CGM data and liver imaging measures, a combination rarely available in population-based studies. Additionally, our multivariable regression models were adjusted for extensive sets of confounders, minimizing the potential for residual confounding. However, VCTE was performed approximately 3-4 years prior to CGM data collection, therefore, it is possible that individuals with hepatic features progressed or regressed during the period. Additionally, the low prevalence of hepatic fibrosis in our study sample warrants cautious interpretations of these results. Lastly, FHS participants mainly comprise of White individuals with European ancestry who mainly reside in one geographical location, which may limit the generalizability of our results. Additional studies with larger, more diverse samples are needed to investigate cross-sectional and longitudinal associations of glycemic traits and hepatic features.

In summary, dynamic measures of glycemic, including 2-h PPG following a mixed meal challenge and CGM-derived summary measures, captured associations with hepatic features beyond traditional measures (HbA1c) among individuals without T2D. Future studies are needed to investigate the longitudinal associations of CGM measures of glycemic burden and variability with MASLD development and progression, in addition to assessing the clinical utility of CGM in MASLD subtyping.

## Data Availability

All data produced in the present study are available upon reasonable request to the authors.

## Acknowledgements

We thank FHS participants for their continued support of the study. BB, NS, HL, MN, JN, DWS, MTL, MEW, and NLS designed research, contributed to discussion, and reviewed/edited the manuscript; BB, MEW, and NLS conducted research, supervised all analyses and writing; BB analyzed data; BB wrote the manuscript.

## Abbreviations

FPG: Fasting plasma glucose
HbA1c: Hemoglobin A1c
T2D: Type 2 diabetes
CGM: Continuous glucose monitoring
2-h PPG: 2-h postprandial glucose
MMTT: Mixed meal tolerance test
MASLD: Metabolic dysfunction-associated steatotic liver disease
CVD: Cardiovascular disease
VCTE: Vibration-controlled transient elastography
CAP: Controlled attenuation parameter
LSM: Liver stiffness measurement
CV: Coefficient of variation
MAGE: Mean amplitude of glycemic excursions
CONGA-1: Continuous overall net glycemic action over 1-hour
MODD: Mean of daily differences
GRADE: Glycemic risk assessment diabetes equation
LBGI: Low blood glucose index
HBGI: High blood glucose index
OGTT: Oral glucose tolerance test

---

**Figure S1.**
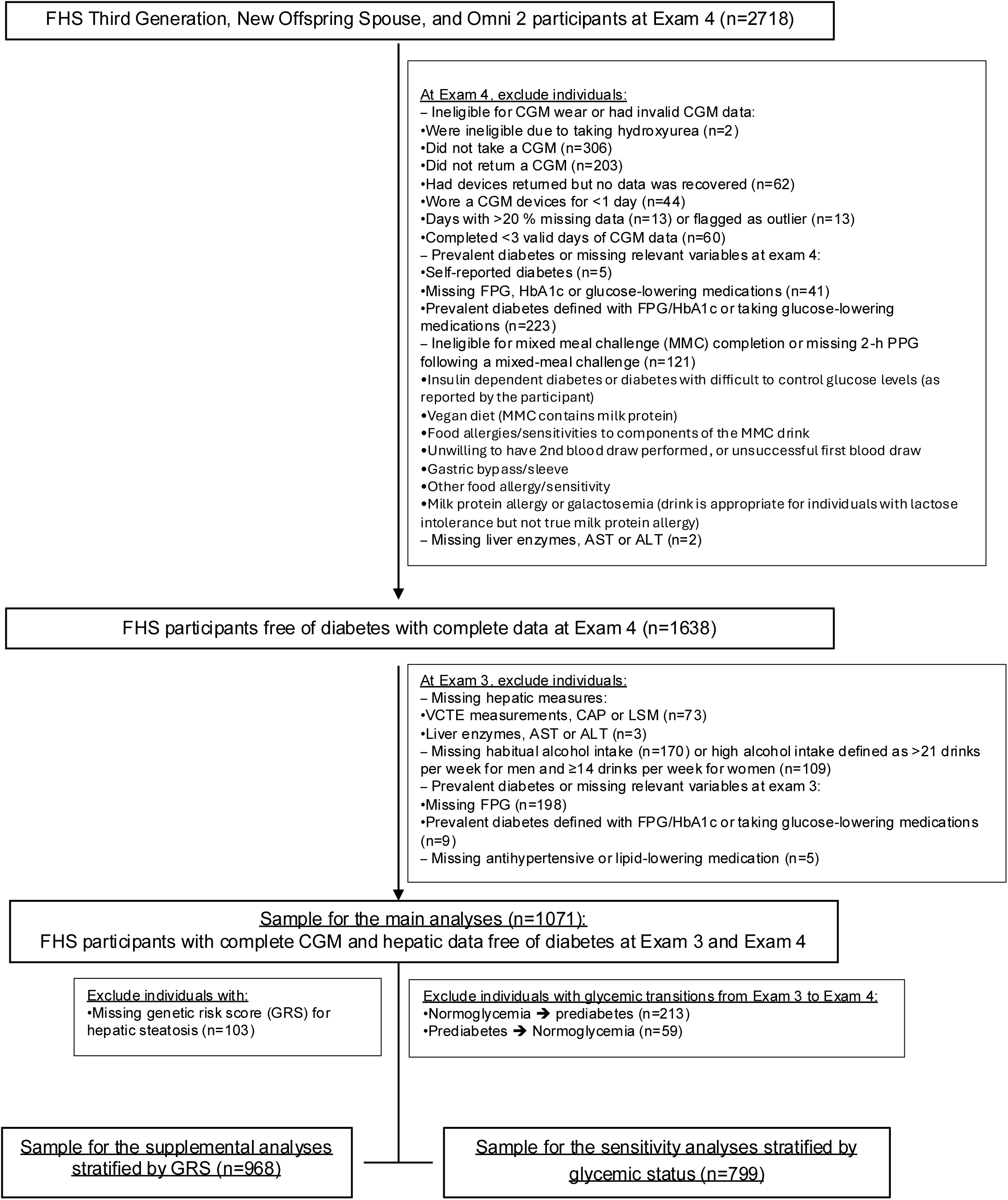
Flowchart of FHS participant exclusions across Exam 3 and Exam 4.

**Figure S2.**
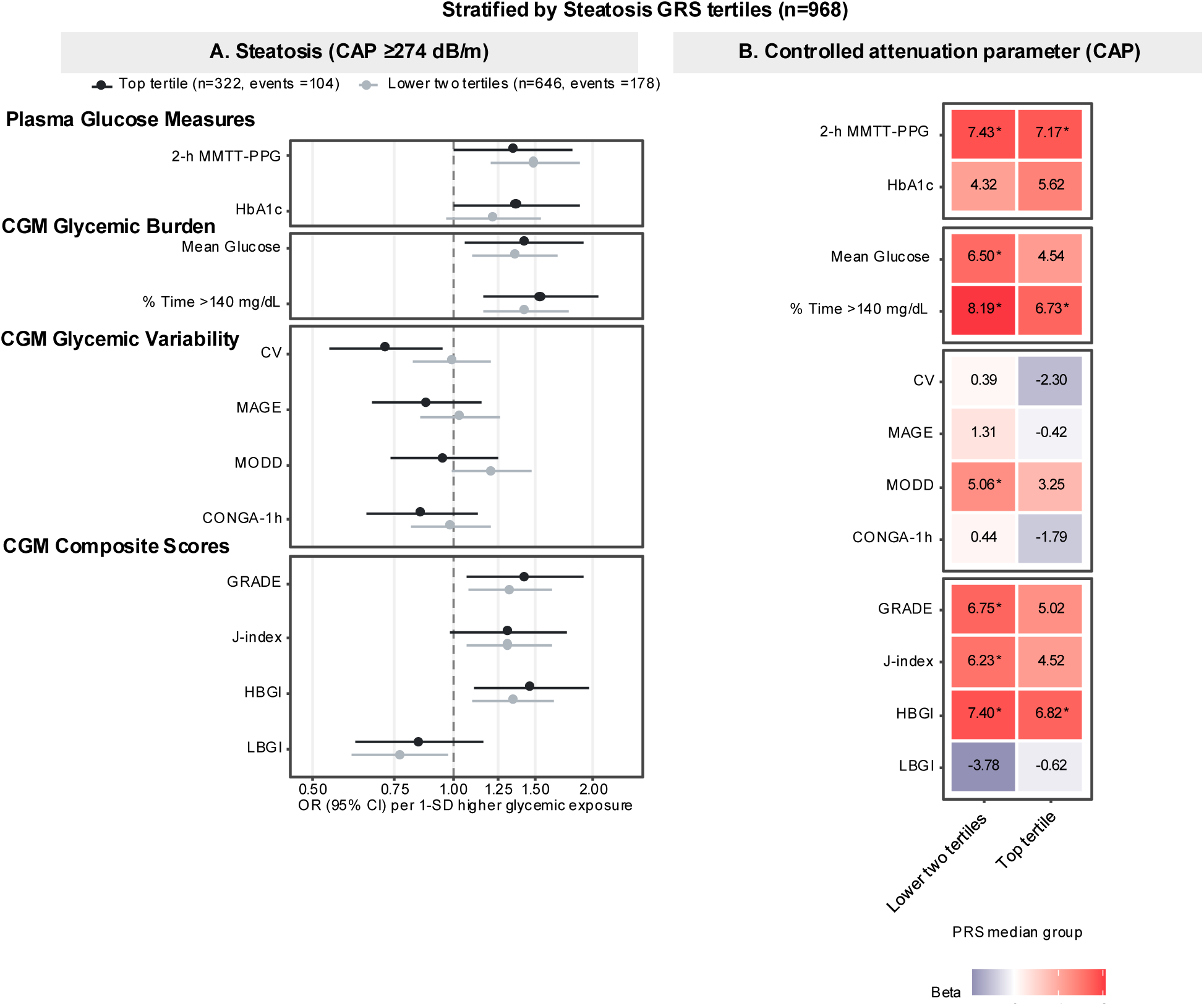
Multivariable linear and logistic regression models depicting the associations between glycemic traits, CAP, and hepatic steatosis, stratified by top tertile versus two lower tertiles of GRS (n=968). Model was adjusted for age, sex, smoking, lipid-lowering medication, alcohol intake, BMI, AST:ALT ratio at Exam 3.

**Figure S3.**
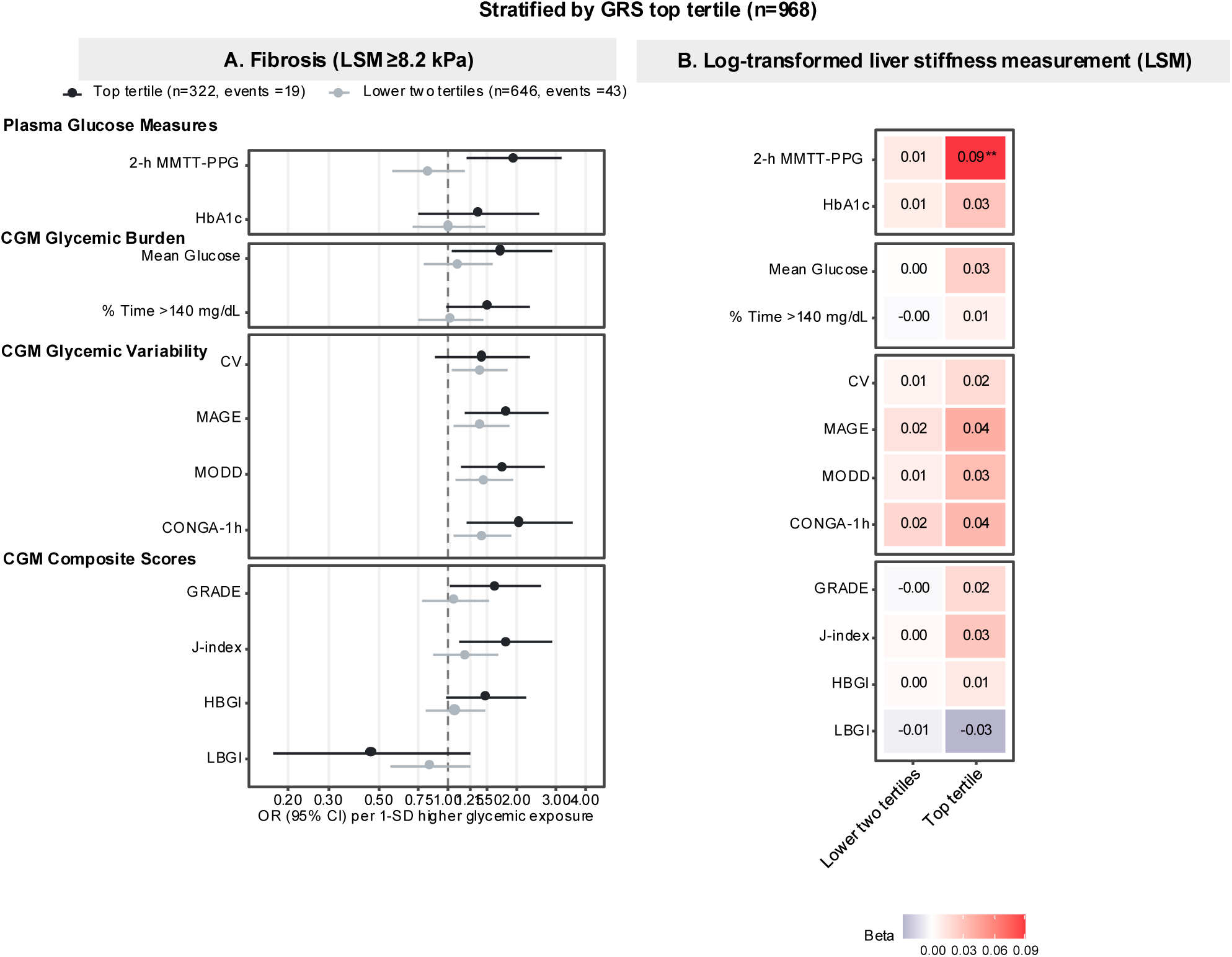
Multivariable linear and logistic regression models depicting the associations between glycemic traits, log-LSM, and hepatic fibrosis, stratified by top tertile versus two lower tertiles of GRS (n=968). Model was adjusted for age, sex, smoking, lipid-lowering medication, alcohol intake, BMI, AST:ALT ratio at Exam 3.

**Figure S4.**
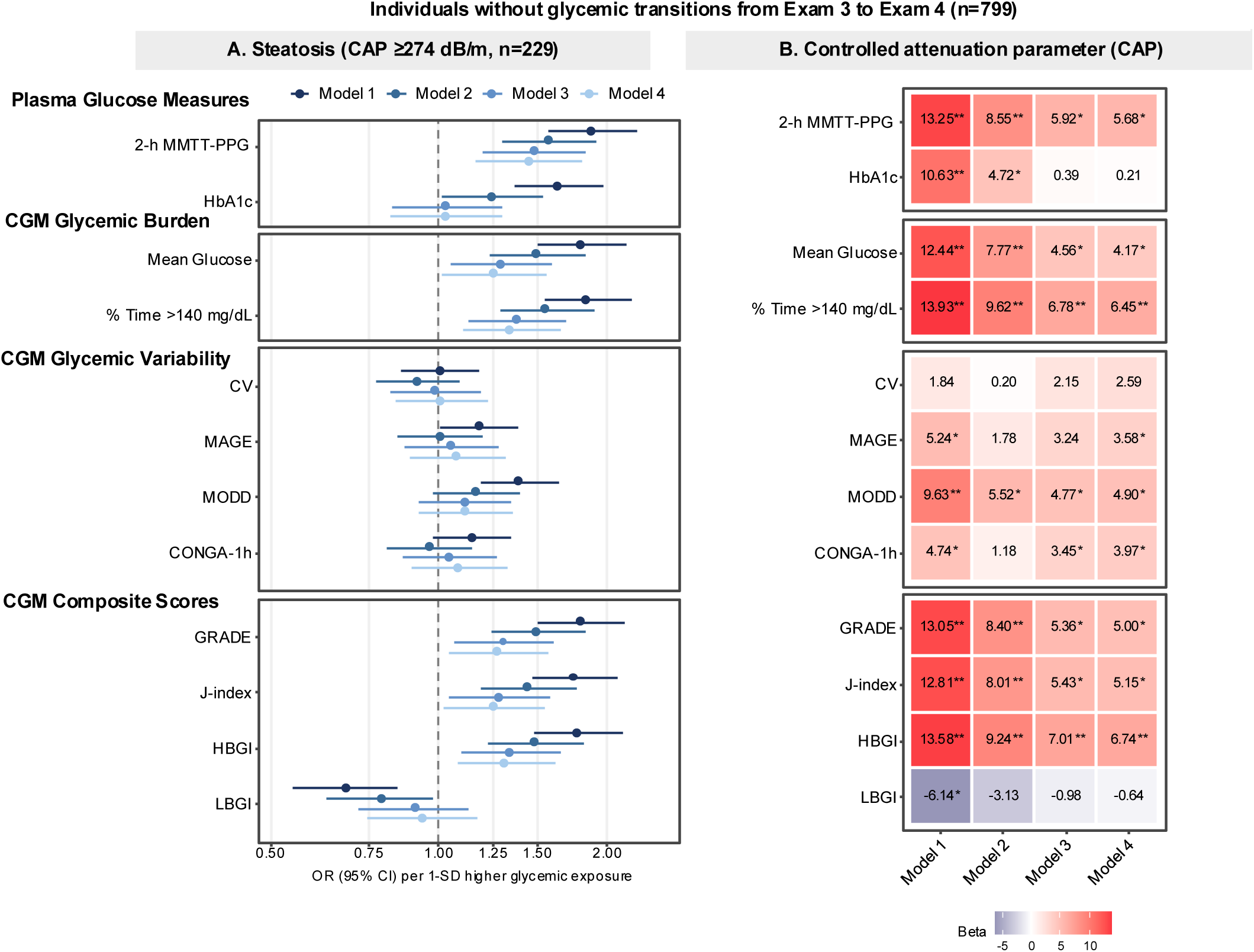
Multivariable linear and logistic regression models depicting the associations between glycemic traits, log-LSM, and hepatic fibrosis, stratified by top tertile versus two lower tertiles of GRS (n=968). Model was adjusted for age, sex, smoking, lipid-lowering medication, alcohol intake, BMI, AST:ALT ratio at Exam 3.

**Figure S5.**
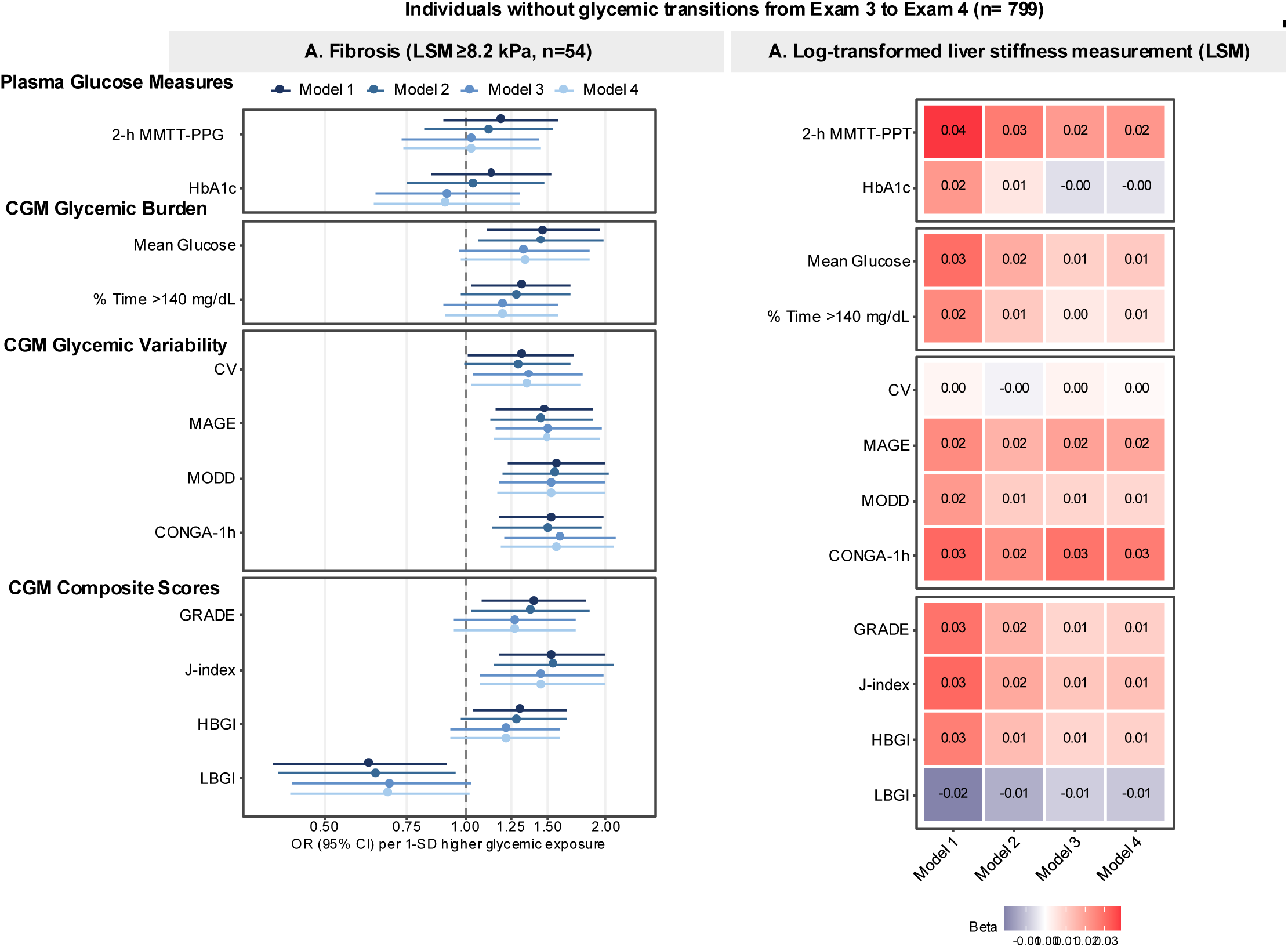
Multivariable linear and logistic regression models depicting the associations between glycemic traits, log-LSM, and hepatic fibrosis, after excluding individuals with glycemic transitions from Exam 3 to Exam 4. Model was adjusted for age, sex, smoking, lipid-lowering medication, alcohol intake at Exam 3, FPG, BMI, and AST:ALT ratio at Exam 3

**Figure S6.**
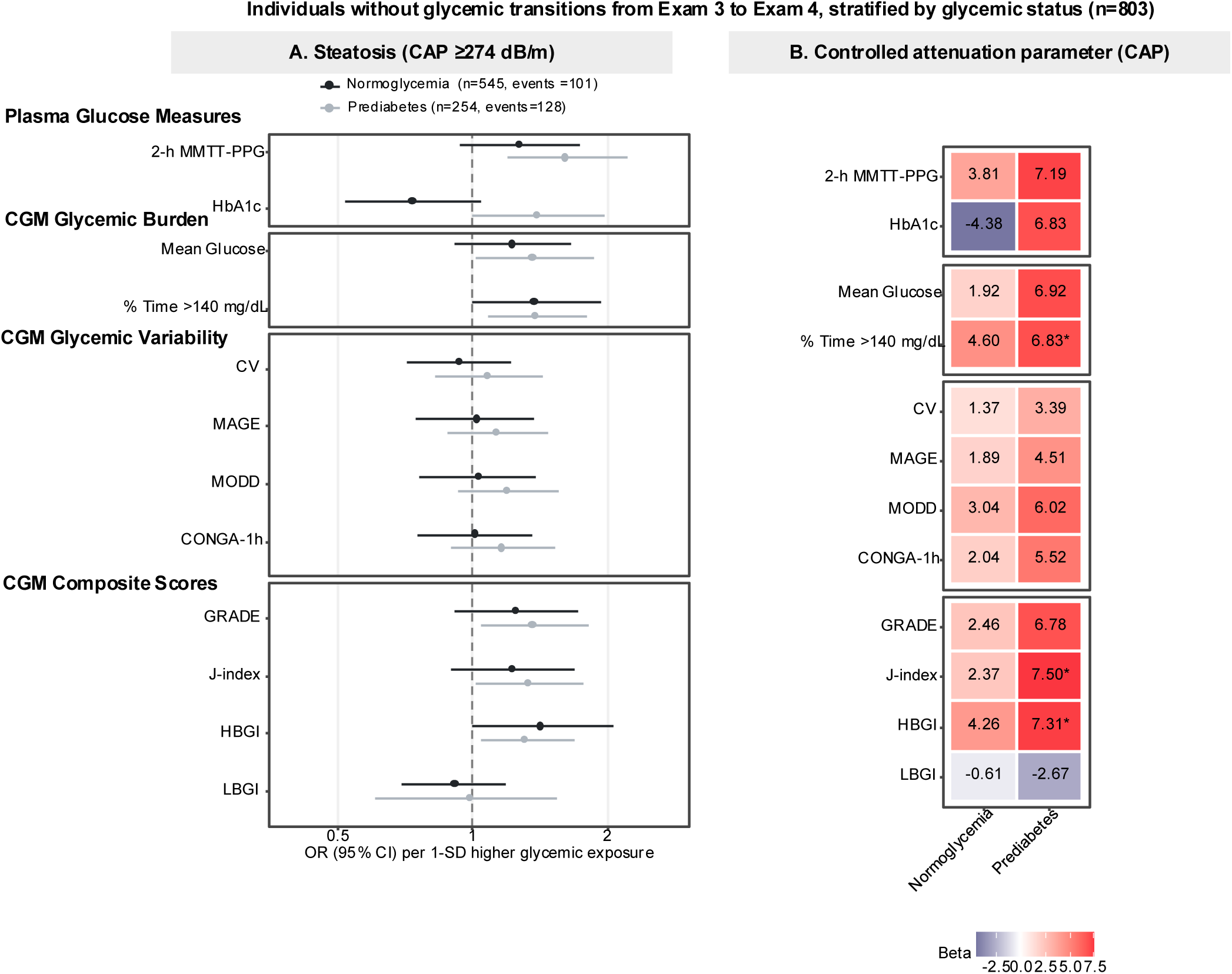
Multivariable linear and logistic regression models depicting the associations between glycemic traits, CAP, and hepatic steatosis among individuals without glycemic transitions, stratified by stable normoglycemia and prediabetes. Model was adjusted for age, sex, smoking, lipid-lowering medication, alcohol intake, BMI, AST:ALT ratio at Exam 3.

**Figure S7.**
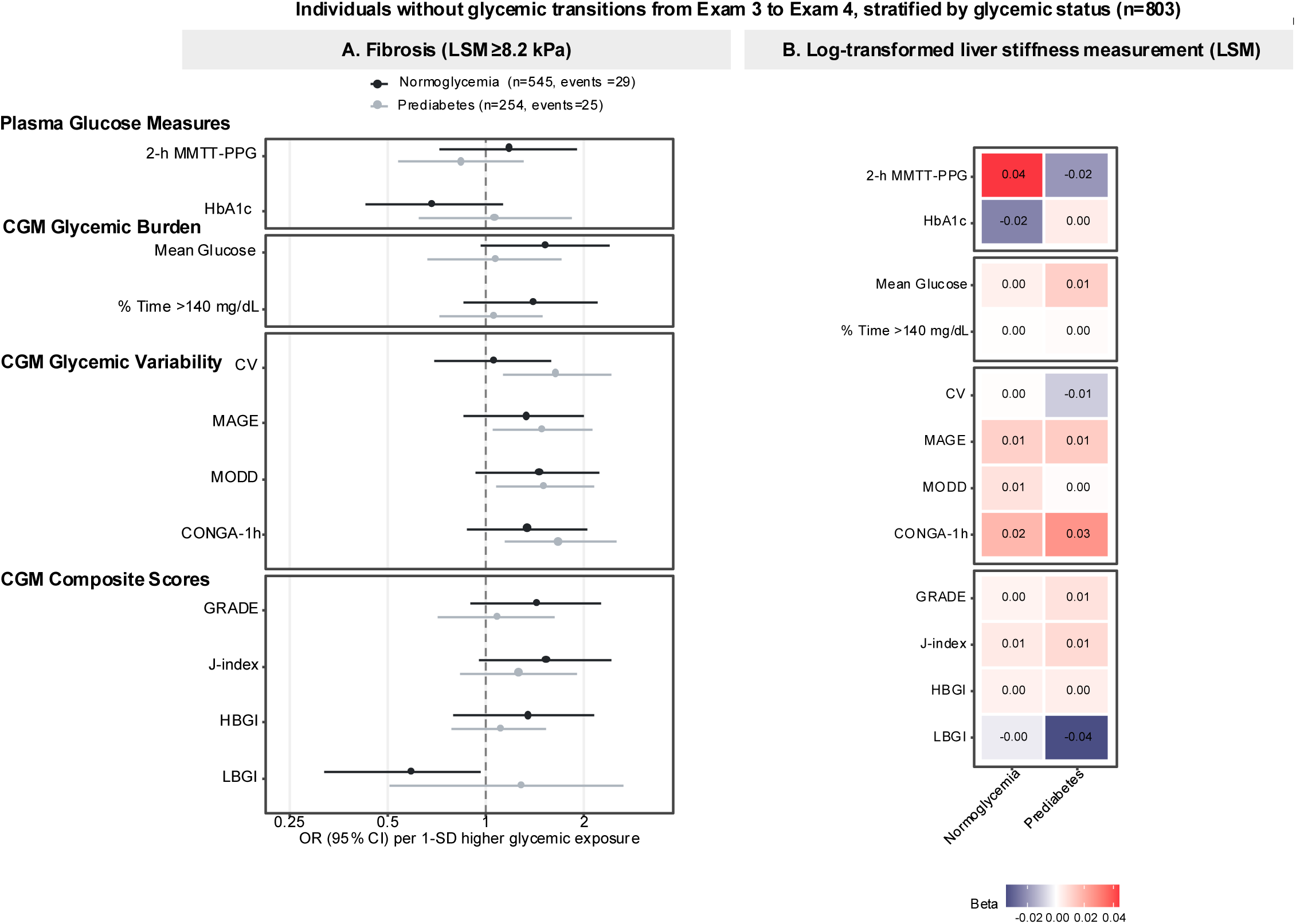
Multivariable linear and logistic regression models depicting the associations between glycemic traits, log-LSM, and hepatic fibrosis among individuals without glycemic transitions, stratified by stable normoglycemia and prediabetes. Model adjusted for age, sex, smoking, lipid-lowering medication, alcohol intake, BMI, AST:ALT ratio at Exam 3.

**Figure S8.**
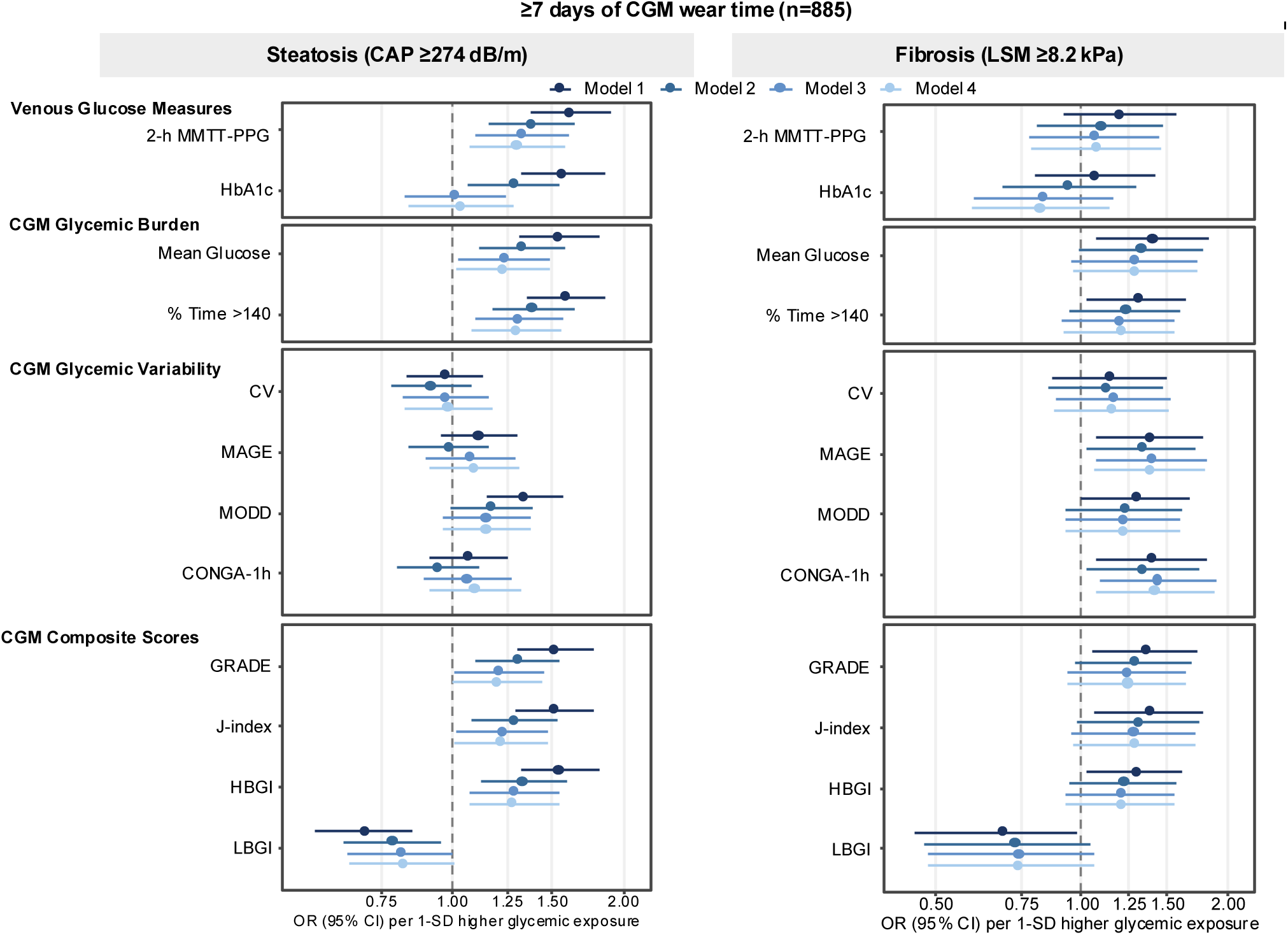
Multivariable logistic regression models depicting the associations between glycemic traits, hepatic steatosis and fibrosis,excluding those with <7 days of CGM wear time. Model 1 was adjusted for age, sex, smoking, lipid-lowering medication, alcohol intake, model 2 included model 1 with additional adjustment for FPG, model 3 included model 2 with additional adjustment for BMI, model 4: model 3 with additional adjustment for additional adjustment for AST:ALT ratio at Exam 3.

