## Supplementary Tables for "Dynamic Measures of Glycemia in Relation to Hepatic Features in Adults Without Diabetes"

| **Table S1.** Details of CGM-derived summary measures. Mean glucose is the average glucose level during the entire wear time, CV is a standardized measure of variability, J-index is a measure of glycemic control that integrates both mean glucose and standard deviation.13,14 MAGE and CONGA-1 are both measures of within-day variability, while MODD is a measure of between-day variability.15,16 GRADE is a composite measure, capturing derivations from an ideal glucose level (90 mg/dL), therefore accounting for both hypo- and hyperglycemia.17 LBGI and HBGI quantify hypo and hyperglycemia, respectively. | |
| --- | --- |
| Mean glucose (mg/dL) | The average interstitial glucose levels during the entire wear time. |
| Time >140 mg/dL (%) | The proportion of monitored time in which interstitial glucose values are above 140 mg/dL.^1^ |
| CV (%) |  |
| MAGE (mg/dL) | A measure of within-day glycemic variability, averaging the magnitude of excursions between consecutive peaks (highest points) and nadirs (lowest points) during the wear time, while only counting the changes in glucose levels that are greater than 1 SD of the glucose for a 24-hour period.^2^ |
| CONGA-1h (mg/dL) | Measure of within-day glycemic variability, representing the differences between consecutive glucose readings taken at 1-hour intervals across the wear time.^3^ |
| MODD (mg/dL) | A measure of between-day glycemic variability, representing the average absolute differences in glucose levels at the same time point on consecutive days.^4^ |
| J-index | An index combining mean and standard deviation (SD) of interstitial glucose to measure the quality of glycemic control:^5^  J-index = 0.001 × (mean + SD)^2^ |
| GRADE | A composite metric that assesses overall glycemic risk by applying a nonlinear weighting function to glucose values, emphasizing clinically important hypo- and hyperglycemic excursions.^6^ |
| HBGI | Quantifies the risk of hyperglycemia.^7^ |
| LBGI | Quantifies the risk of hypoglycemia.^7^ |

**Abbreviations:** CV, coefficient of variation; MAGE, mean amplitude of glycemic excursions; CONGA-1, continuous overall net glycemic action (1-hour); MODD, mean of daily differences; GRADE, Glycemic Risk Assessment Diabetes Equation; HBGI, high blood glucose index; LBGI: low blood glucose index.

| **Table S2.** Participant characteristics according to hepatic steatosis. | | | |
| --- | --- | --- | --- |
|  | **Overall**  n = 1,071 | **No steatosis**  n = 757 | **Steatosis**  n = 314 |
| **Exam 4 demographics and clinical characteristics** | | | |
| Age, years | 60.2 (8.4) | 59.7 (8.8) | 61.4 (7.2) |
| Female | 625 (58.4%) | 483 (63.8%) | 142 (45.2%) |
| Body mass index, kg/m² | 28.1 (5.5) | 26.6 (4.7) | 31.8 (5.5) |
| Triglycerides, mg/dL | 87.0 (66.0, 120.0) | 81.0 (63.0, 111.0) | 107.5 (80.0, 149.0) |
| Total cholesterol, mg/dL | 185.0 (162.0, 208.0) | 187.0 (166.0, 210.0) | 180.0 (156.0, 204.0) |
| HDL cholesterol, mg/dL | 60.6 (17.9) | 63.7 (18.0) | 53.2 (15.3) |
| Blood pressure medication | 269 (25.1%) | 146 (19.3%) | 123 (39.2%) |
| Lipid-lowering medication | 331 (30.9%) | 201 (26.6%) | 130 (41.4%) |
| **Venous glycemic measures** | | | |
| FPG, mg/dL | 97.5 (9.4) | 95.7 (8.7) | 101.7 (9.8) |
| HbA1c, % | 5.3 (0.3) | 5.3 (0.3) | 5.5 (0.4) |
| 2-hour PPG, mg/dL | 108.1 (21.9) | 104.4 (20.8) | 116.8 (22.2) |
| **CGM summary measures** | | | |
| CGM wear time, days | 8.2 (1.4) | 8.3 (1.4) | 8.1 (1.5) |
| Mean CGM glucose, mg/dL | 118.4 (13.7) | 116.2 (12.2) | 123.6 (15.5) |
| time >140 mg/dL, % | 15.2 (15.4) | 12.5 (11.9) | 21.7 (20.1) |
| Coefficient of variation, % | 16.2 (3.4) | 16.2 (3.4) | 16.3 (3.5) |
| MAGE | 48.7 (13.2) | 47.8 (12.8) | 50.7 (13.8) |
| MODD | 17.4 (4.3) | 16.9 (3.8) | 18.6 (5.1) |
| CONGA-1 | 21.1 (5.1) | 20.9 (5.1) | 21.7 (5.1) |
| J-index | 18.5 (15.9, 21.5) | 17.9 (15.5, 20.8) | 19.9 (17.2, 23.4) |
| GRADE | 2.5 (1.6, 3.5) | 2.3 (1.6, 3.2) | 2.9 (2.0, 4.1) |
| LBGI | 0.3 (0.1, 0.6) | 0.3 (0.1, 0.7) | 0.2 (0.1, 0.5) |
| HBGI | 0.6 (0.3, 1.1) | 0.5 (0.3, 1.0) | 0.8 (0.4, 1.5) |
| **Exam 3 hepatic phenotypes and related covariates** | | | |
| AST, U/L | 21.0 (18.0, 25.0) | 21.0 (18.0, 25.0) | 22.0 (19.0, 27.0) |
| ALT, U/L | 20.0 (15.0, 27.0) | 18.0 (15.0, 23.0) | 25.0 (18.0, 33.0) |
| CAP, dB/m | 248.4 (51.1) | 222.4 (31.9) | 310.9 (30.2) |
| LSM, kPa | 4.9 (4.0, 6.1) | 4.8 (4.0, 6.0) | 5.5 (4.3, 6.6) |
| Alcohol intake, g/day | / | 7.5 (2.3, 14.7) | 5.7 (1.5, 13.9) |
| Characteristics presented as Mean (SD) or Median (Q1, Q3) for continuous and n (%) for binary variables. | | | |
| **Abbreviations:** FPG, fasting plasma glucose; 2-h PPG, 2-hour post–MMTT glucose; AST, aspartate aminotransferase; ALT, alanine aminotransferase; CAP, controlled attenuation parameter; LSM, liver stiffness measurement; CV, coefficient of variation; MAGE, mean amplitude of glycemic excursions; CONGA-1, continuous overall net glycemic action (1-hour); MODD, mean of daily differences; GRADE, Glycemic Risk Assessment Diabetes Equation; HBGI, high blood glucose index; LBGI, low blood glucose index. | | | |

| **Table S3.** Multivariable logistic regression models depicting the associations between standardized glycemic traits and hepatic steatosis (n=1071, steatosis=314) | | | | | | | | |
| --- | --- | --- | --- | --- | --- | --- | --- | --- |
|  | **Model 1** | | **Model 2** | | **Model 3** | | **Model 4** | |
| **Exposure** | **OR (95% CI)** | **p-value** | **OR (95% CI)** | **p-value** | **OR (95% CI)** | **p-value** | **OR (95% CI)** | **p-value** |
| **Venous Glucose Measures** | | | | | | | | |
| 2-h MMTT-PPG | 1.73 (1.49, 2.01) | <0.001 | 1.48 (1.26, 1.74) | <0.001 | 1.39 (1.17, 1.65) | <0.001 | 1.36 (1.14, 1.63) | <0.01 |
| HbA1c | 1.51 (1.30, 1.76) | <0.001 | 1.23 (1.05, 1.46) | 0.02 | 1.01 (0.84, 1.22) | 0.91 | 1.04 (0.86, 1.25) | 0.80 |
| **CGM Glycemic Burden** | | | | | | | | |
| Mean Glucose | 1.57 (1.36, 1.82) | <0.001 | 1.34 (1.15, 1.57) | <0.001 | 1.26 (1.06, 1.49) | 0.02 | 1.25 (1.05, 1.48) | 0.02 |
| % Time >140 | 1.60 (1.39, 1.85) | <0.001 | 1.39 (1.19, 1.62) | <0.001 | 1.32 (1.13, 1.55) | <0.01 | 1.31 (1.12, 1.55) | <0.01 |
| **CGM Glycemic Variability** | | | | | | | | |
| CV | 0.99 (0.86, 1.14) | 0.91 | 0.93 (0.80, 1.07) | 0.38 | 0.97 (0.83, 1.14) | 0.82 | 0.98 (0.84, 1.15) | 0.91 |
| MAGE | 1.14 (0.99, 1.30) | 0.10 | 1.00 (0.86, 1.16) | 0.99 | 1.07 (0.91, 1.26) | 0.50 | 1.08 (0.92, 1.28) | 0.41 |
| MODD | 1.32 (1.15, 1.52) | <0.001 | 1.15 (1.00, 1.33) | 0.09 | 1.13 (0.96, 1.32) | 0.19 | 1.12 (0.96, 1.32) | 0.21 |
| CONGA-1h | 1.09 (0.95, 1.25) | 0.28 | 0.96 (0.83, 1.11) | 0.64 | 1.07 (0.91, 1.26) | 0.50 | 1.10 (0.93, 1.29) | 0.36 |
| **CGM Composite Scores** | | | | | | | | |
| GRADE | 1.55 (1.34, 1.78) | <0.001 | 1.32 (1.13, 1.54) | <0.01 | 1.24 (1.05, 1.46) | 0.02 | 1.23 (1.04, 1.46) | 0.02 |
| J-index | 1.53 (1.33, 1.76) | <0.001 | 1.30 (1.11, 1.52) | <0.01 | 1.24 (1.05, 1.46) | 0.02 | 1.23 (1.04, 1.46) | 0.03 |
| HBGI | 1.54 (1.34, 1.78) | <0.001 | 1.32 (1.14, 1.55) | <0.01 | 1.28 (1.09, 1.51) | <0.01 | 1.27 (1.09, 1.50) | <0.01 |
| LBGI | 0.71 (0.59, 0.84) | <0.001 | 0.79 (0.66, 0.94) | 0.02 | 0.83 (0.69, 1.00) | 0.09 | 0.84 (0.69, 1.01) | 0.11 |
| Model 1: Age, Sex, Smoking, Lipid-lowering medication, Alcohol intake  Model 2: Age, Sex, Smoking, Lipid-lowering medication, Alcohol intake, Fasting glucose  Model 3: Age, Sex, Smoking, Lipid-lowering medication, Alcohol intake, Fasting glucose, BMI  Model 4: Age, Sex, Smoking, Lipid-lowering medication, Alcohol intake, Fasting glucose, BMI, AST:ALT ratio at Exam 3 | | | | | | | | |

| **Table S4.** Multivariable linear regression models depicting the associations between standardized glycemic traits and controlled attenuation parameter (CAP). | | | | | | | | |
| --- | --- | --- | --- | --- | --- | --- | --- | --- |
|  | **Model 1** | | **Model 2** | | **Model 3** | | **Model 4** | |
| **Exposure** | **Beta (95% CI)** | **p-value** | **Beta (95% CI)** | **p-value** | **Beta (95% CI)** | **p-value** | **Beta (95% CI)** | **p-value** |
| **Plasma Glucose Measures** | | | | | | | | |
| 2-h MMTT-PPG | 12.04 (9.02, 15.06) | <0.001 | 7.85 (4.55, 11.14) | <0.001 | 5.18 (2.17, 8.19) | <0.01 | 4.86 (1.90, 7.83) | <0.01 |
| HbA1c | 8.83 (5.74, 11.92) | <0.001 | 3.93 (0.59, 7.27) | 0.03 | -0.80 (-3.87, 2.28) | 0.65 | -0.63 (-3.66, 2.40) | 0.71 |
| **CGM Glycemic Burden** | | | | | | | | |
| Mean Glucose | 9.64 (6.57, 12.71) | <0.001 | 5.50 (2.26, 8.74) | <0.01 | 3.55 (0.61, 6.49) | 0.03 | 3.36 (0.46, 6.26) | 0.03 |
| % Time >140 mg/dL | 10.79 (7.74, 13.83) | <0.001 | 6.97 (3.77, 10.17) | <0.001 | 5.29 (2.39, 8.19) | <0.01 | 5.04 (2.18, 7.90) | <0.01 |
| **CGM Glycemic Variability** | | | | | | | | |
| CV | 1.12 (-1.89, 4.13) | 0.52 | -0.03 (-2.96, 2.89) | 0.98 | 1.69 (-0.96, 4.34) | 0.25 | 1.99 (-0.62, 4.60) | 0.17 |
| MAGE | 3.74 (0.70, 6.78) | 0.03 | 0.93 (-2.10, 3.95) | 0.60 | 2.74 (0.01, 5.48) | 0.07 | 3.02 (0.33, 5.72) | 0.04 |
| MODD | 8.08 (5.08, 11.08) | <0.001 | 4.85 (1.80, 7.90) | <0.01 | 4.35 (1.60, 7.11) | <0.01 | 4.38 (1.67, 7.09) | <0.01 |
| CONGA-1h | 2.65 (-0.38, 5.67) | 0.11 | -0.13 (-3.14, 2.87) | 0.95 | 2.67 (-0.06, 5.40) | 0.07 | 3.14 (0.45, 5.83) | 0.03 |
| **CGM Composite Scores** | | | | | | | | |
| GRADE | 9.91 (6.85, 12.97) | <0.001 | 5.72 (2.48, 8.97) | <0.01 | 3.89 (0.94, 6.84) | 0.02 | 3.70 (0.80, 6.60) | 0.02 |
| J-index | 9.77 (6.70, 12.84) | <0.001 | 5.50 (2.24, 8.76) | <0.01 | 4.10 (1.15, 7.05) | 0.01 | 3.96 (1.05, 6.87) | 0.01 |
| HBGI | 10.39 (7.37, 13.41) | <0.001 | 6.49 (3.30, 9.67) | <0.001 | 5.30 (2.42, 8.18) | <0.01 | 5.13 (2.29, 7.97) | <0.01 |
| LBGI | -5.56 (-8.60, -2.52) | <0.01 | -2.89 (-5.91, 0.14) | 0.08 | -1.49 (-4.23, 1.25) | 0.34 | -1.30 (-4.00, 1.40) | 0.39 |
| Model 1: Age, Sex, Smoking, Lipid-lowering medication, Alcohol intake  Model 2: Age, Sex, Smoking, Lipid-lowering medication, Alcohol intake, Fasting glucose  Model 3: Age, Sex, Smoking, Lipid-lowering medication, Alcohol intake, Fasting glucose, BMI  Model 4: Age, Sex, Smoking, Lipid-lowering medication, Alcohol intake, Fasting glucose, BMI, AST:ALT ratio at Exam 3 | | | | | | | | |

| **Table S5.** Multivariable logistic regression models depicting the associations between standardized glycemic traits and hepatic fibrosis (n=1071, fibrosis= 71) | | | | | | | | |
| --- | --- | --- | --- | --- | --- | --- | --- | --- |
|  | **Model 1** | | **Model 2** | | **Model 3** | | **Model 4** | |
| **Exposure** | **OR (95% CI)** | **p-value** | **OR (95% CI)** | **p-value** | **OR (95% CI)** | **p-value** | **OR (95% CI)** | **p-value** |
| **Plasma Glucose Measures** | | | | | | | | |
| 2-h Post-prandial Glucose | 1.15 (0.90, 1.46) | 0.30 | 1.08 (0.82, 1.42) | 0.64 | 1.02 (0.76, 1.35) | 0.89 | 1.03 (0.77, 1.36) | 0.88 |
| HbA1c | 1.11 (0.86, 1.44) | 0.46 | 1.04 (0.78, 1.38) | 0.83 | 0.93 (0.70, 1.24) | 0.66 | 0.92 (0.69, 1.24) | 0.65 |
| **CGM Glycemic Burden** | | | | | | | | |
| Mean Glucose | 1.44 (1.13, 1.81) | <0.01 | 1.42 (1.09, 1.84) | 0.02 | 1.37 (1.05, 1.78) | 0.03 | 1.37 (1.05, 1.78) | 0.03 |
| % Time >140 | 1.32 (1.06, 1.60) | 0.02 | 1.29 (1.02, 1.61) | 0.04 | 1.25 (0.99, 1.56) | 0.07 | 1.25 (0.99, 1.56) | 0.07 |
| **CGM Glycemic Variability** | | | | | | | | |
| CV | 1.30 (1.03, 1.62) | 0.04 | 1.28 (1.01, 1.61) | 0.05 | 1.32 (1.05, 1.66) | 0.03 | 1.32 (1.04, 1.66) | 0.03 |
| MAGE | 1.50 (1.21, 1.85) | <0.01 | 1.48 (1.18, 1.83) | <0.01 | 1.54 (1.23, 1.92) | <0.01 | 1.54 (1.22, 1.92) | <0.01 |
| MODD | 1.49 (1.20, 1.84) | <0.01 | 1.47 (1.17, 1.84) | <0.01 | 1.46 (1.16, 1.83) | <0.01 | 1.46 (1.16, 1.83) | <0.01 |
| CONGA-1h | 1.51 (1.20, 1.90) | <0.01 | 1.49 (1.17, 1.88) | <0.01 | 1.60 (1.25, 2.03) | <0.01 | 1.59 (1.24, 2.03) | <0.01 |
| **CGM Composite Scores** | | | | | | | | |
| GRADE | 1.38 (1.10, 1.70) | 0.01 | 1.36 (1.06, 1.72) | 0.03 | 1.31 (1.02, 1.67) | 0.04 | 1.32 (1.02, 1.68) | 0.04 |
| J-index | 1.47 (1.17, 1.83) | <0.01 | 1.47 (1.14, 1.88) | <0.01 | 1.43 (1.11, 1.84) | 0.01 | 1.44 (1.11, 1.85) | 0.01 |
| HBGI | 1.32 (1.08, 1.59) | 0.01 | 1.29 (1.04, 1.59) | 0.03 | 1.27 (1.02, 1.57) | 0.04 | 1.28 (1.02, 1.57) | 0.04 |
| LBGI | 0.66 (0.44, 0.92) | 0.04 | 0.68 (0.45, 0.95) | 0.05 | 0.69 (0.46, 0.98) | 0.07 | 0.69 (0.46, 0.97) | 0.07 |
| Model 1: Age, Sex, Smoking, Lipid-lowering medication, Alcohol intake  Model 2: Age, Sex, Smoking, Lipid-lowering medication, Alcohol intake, Fasting glucose  Model 3: Age, Sex, Smoking, Lipid-lowering medication, Alcohol intake, Fasting glucose, BMI,  Model 4: Age, Sex, Smoking, Lipid-lowering medication, Alcohol intake, Fasting glucose, BMI, AST:ALT ratio at Exam 3 | | | | | | | | |

| **Table S6.** Multivariable linear regression models depicting the associations between standardized glycemic traits and log-transformed liver stiffness measurement (log-LSM). | | | | | | | | |
| --- | --- | --- | --- | --- | --- | --- | --- | --- |
|  | **Model 1** | | **Model 2** | | **Model 3** | | **Model 4** | |
| **Exposure** | **Beta (95% CI)** | **p-value** | **Beta (95% CI)** | **p-value** | **Beta (95% CI)** | **p-value** | **Beta (95% CI)** | **p-value** |
| **Venous Glucose Measures** |  |  |  |  |  |  |  |  |
| 2-h Post-prandial Glucose | 0.04 (0.02, 0.06) | 0.02 | 0.03 (0.01, 0.05) | 0.08 | 0.02 (0.00, 0.05) | 0.13 | 0.02 (0.00, 0.05) | 0.13 |
| HbA1c | 0.02 (0.00, 0.05) | 0.13 | 0.01 (-0.01, 0.03) | 0.48 | -0.00 (-0.03, 0.02) | 0.97 | -0.00 (-0.03, 0.02) | 0.97 |
| **CGM Glycemic Burden** |  |  |  |  |  |  |  |  |
| Mean Glucose | 0.03 (0.01, 0.05) | 0.08 | 0.02 (-0.01, 0.04) | 0.28 | 0.01 (-0.01, 0.04) | 0.35 | 0.01 (-0.01, 0.04) | 0.35 |
| % Time >140 mg/dL | 0.02 (0.00, 0.05) | 0.13 | 0.01 (-0.01, 0.04) | 0.39 | 0.01 (-0.01, 0.03) | 0.50 | 0.01 (-0.01, 0.03) | 0.50 |
| **CGM Glycemic Variability** |  |  |  |  |  |  |  |  |
| CV | 0.01 (-0.01, 0.03) | 0.50 | 0.00 (-0.02, 0.03) | 0.68 | 0.01 (-0.01, 0.03) | 0.48 | 0.01 (-0.01, 0.03) | 0.49 |
| MAGE | 0.03 (0.01, 0.05) | 0.08 | 0.02 (-0.00, 0.04) | 0.14 | 0.03 (0.00, 0.05) | 0.10 | 0.03 (0.00, 0.05) | 0.10 |
| MODD | 0.02 (0.00, 0.05) | 0.11 | 0.02 (-0.01, 0.04) | 0.32 | 0.01 (-0.01, 0.04) | 0.32 | 0.01 (-0.01, 0.04) | 0.32 |
| CONGA-1h | 0.03 (0.01, 0.05) | 0.08 | 0.02 (0.00, 0.04) | 0.13 | 0.03 (0.01, 0.05) | 0.08 | 0.03 (0.01, 0.05) | 0.08 |
| **CGM Composite Scores** |  |  |  |  |  |  |  |  |
| GRADE | 0.03 (0.00, 0.05) | 0.08 | 0.02 (-0.01, 0.04) | 0.32 | 0.01 (-0.01, 0.04) | 0.41 | 0.01 (-0.01, 0.04) | 0.41 |
| J-index | 0.03 (0.01, 0.05) | 0.08 | 0.02 (-0.00, 0.04) | 0.26 | 0.02 (-0.01, 0.04) | 0.32 | 0.02 (-0.01, 0.04) | 0.32 |
| HBGI | 0.03 (0.01, 0.05) | 0.08 | 0.02 (-0.01, 0.04) | 0.31 | 0.01 (-0.01, 0.04) | 0.33 | 0.01 (-0.01, 0.04) | 0.33 |
| LBGI | -0.02 (-0.04, -0.00) | 0.13 | -0.02 (-0.04, 0.01) | 0.32 | -0.01 (-0.03, 0.01) | 0.35 | -0.01 (-0.03, 0.01) | 0.35 |
